# Comparison of *TP53* Mutations in Myelodysplasia and Acute Leukemia Suggests Divergent Roles in Initiation and Progression

**DOI:** 10.1101/2023.09.04.23295042

**Authors:** Ashwini Jambhekar, Emily E. Ackerman, Berk A. Alpay, Galit Lahav, Scott B. Lovitch

**Affiliations:** Department of Systems Biology, Harvard Medical School, Boston, MA; Ludwig Center at Harvard, Boston, MA; Systems, Synthetic, and Quantitative Biology Program, Harvard University, Cambridge, MA; Department of Organismal and Evolutionary Biology, Harvard University, Cambridge, MA; Department of Pathology, Brigham and Women’s Hospital, Boston, MA

## Abstract

*TP53* mutation predicts adverse prognosis in many cancers, including myeloid neoplasms, but the mechanisms by which specific mutations impact disease biology, and whether they differ between disease categories, remain unknown. We analyzed *TP53* mutations in four myeloid neoplasm subtypes (MDS, AML, AML with myelodysplasia-related changes (AML-MRC), and therapy-related acute myeloid leukemia (tAML)), and identified differences in mutation types, spectrum, and hotspots between disease categories and compared to solid tumors. Missense mutations in the DNA-binding domain were most common across all categories, whereas inactivating mutations and mutations outside the DNA binding domain were more common in AML-MRC compared to MDS. *TP53* mutations in MDS were more likely to retain transcriptional activity, and co-mutation profiles were distinct between disease categories and mutation types. Our findings suggest that mutated *TP53* contributes to initiation and progression of neoplasia via distinct mechanisms, and support the utility of specific identification of *TP53* mutations in myeloid malignancies.

**Statement of Significance:** The distribution and functional consequences of *TP53* mutations differ between hematologic malignancies and solid tumors, and, among myeloid neoplasms, between myelodysplastic syndrome and acute leukemia. These findings suggest distinct biological mechanisms for mutated p53 in hematologic malignancies, specifically in initiation and progression of myeloid neoplasia, that warrant further investigation.

## Introduction

The transcription factor p53, encoded by the *TP53* gene, functions as a tumor suppressor in multiple tissues (1). *TP53* is the most frequently mutated gene in human cancers, with mutation incidence exceeding 50% for many cancer types (2). In myeloid neoplasms, *TP53* mutations occur in approximately 10% of cases (3, 4), and, as for solid tumors, are strong and independent predictors of adverse prognosis, including failure of induction chemotherapy, relapse following hematopoietic stem cell transplantation, and reduced overall survival (5–8). Some studies have reported adverse outcomes only in cases of multiple hits to *TP53* (9), whereas others found even a single hit resulted in poor prognosis compared to *TP53^WT^* neoplasms (10).

Myeloid neoplasms have traditionally been divided into myelodysplastic syndromes (MDS) and acute myeloid leukemia (MDS) by blast enumeration, with >20% blasts in blood or bone marrow generally required for classification as AML; this division persists in the current World Health Organization (WHO) classification (11) and International Consensus Classification (ICC)(12). AML arising via progression from MDS, or following cytotoxic chemotherapy or radiation, have worse prognosis than *de novo* AML and are separately classified; these categories were defined as AML with myelodysplasia-related changes (AML-MRC) and therapy-related AML (t-AML) in the revised 4^th^ edition WHO classification, and are maintained in the 5^th^ edition WHO classification and ICC. Recent studies have demonstrated that the prognostic impact of *TP53* mutation cuts across these traditional classifications, with *TP53*-mutated MDS and AML showing similarly poor prognosis (10, 13); consequently, the WHO 5^th^ edition and ICC both recognize *TP53*-mutated myeloid neoplasia as a specific disease category (11, 12). However, the impact of specific *TP53* mutations in myeloid neoplasms has not been studied.

The mechanisms by which *TP53* mutations contribute to cancer biology are controversial. *TP53* is unusual among tumor suppressor genes in that mutations are strongly biased towards missense mutations, which potentially retain function, rather than inactivating mutations. Mutations are enriched in the DNA-binding domain (DBD): approximately 30% of mutations occur at seven “hotspots”, all within the DBD (14). The vast majority of missense mutations compromise its transcriptional activity, resulting in accumulation of mutant p53 due to its failure to induce MDM2, the E3 ubiquitin ligase that mediates its degradation. However, p53 missense mutations can retain some transcriptional activity: analysis of 2,314 p53 mutants in a reporter assay revealed transcriptional activities from none to wild-type (WT) levels, with most DBD mutations reducing activity below 20% of WT p53 (15). A recent study demonstrated that the distribution of p53 mutational hotspots varies between AML subtypes, suggesting that there may be distinct and context-dependent roles for different mutations type, or for specific mutations (16).

However, there has not been any comprehensive assessment of the distribution of p53 mutations across broader categories of myeloid neoplasia, e.g., MDS versus AML, and current diagnostic criteria for hematologic malignancies do not distinguish between p53 mutation types. With *TP53* mutation assessment is now incorporated into routine diagnostic workup of myeloid neoplasms, it is critical to understand the nature of p53 mutations in these diseases, and their biological functions in modulating disease initiation or progression.

In this study, we analyzed *TP53* mutations in myeloid neoplasms, comparing relative frequencies of mutation types, distributions across the p53 protein, and functional consequences across categories of myeloid neoplasia. We identified significant differences between disease categories in types and locations of mutations, transcriptional capacity, and co-mutation profiles, with the greatest differences occurring between MDS and AML-MRC. These results suggest that *TP53* mutations may play distinct pathogenic roles in different categories of myeloid disease, and that different functional categories of *TP53* mutation may mediate initiation and progression of myeloid neoplasia.

## Results

### *TP53* mutation types and locations vary between myeloid neoplasms

In the analyzed datasets, 6.9-23.5% of cases of myeloid neoplasia were *TP53*-mutant, consistent with previously reported statistics (3). Missense mutations were most prevalent across all disease categories, as expected (16–18). Nevertheless, the spectrum of mutation types varied between neoplasms, with MDS showing significantly more missense mutations and fewer splice-site mutations compared to AML-MRC (**Fig. 1A**). This trend was maintained among cases with multiple *TP53* mutations (**Fig. 1B**) or with VAF > 0.55, a proxy for loss of heterozygosity (**Fig. 1C**). While mutations were strongly enriched in the DBD for all disease categories, as reported (16), DBD enrichment was significantly higher in MDS than in AML or AML-MRC (**Fig. 1D**). A nuclear localization sequence (NLS)-containing region adjacent to the DBD was significantly enriched for mutations in AML-MRC (**Fig. 1D**). In all disease categories, over 80% of DBD mutations were missense, consistent with reports in other cancer types (t-AML was omitted from this and subsequent analyses, due to its small sample size) (17, 18). By contrast, frameshift and nonsense mutations occurred relatively more frequently in other regions, particularly in the NLS (**Fig. 1E**). The first activation domain (AD1) did not contain any nonsense mutations in any disease category (**Fig. 1C**). Overall, these results recapitulated the general trend of enrichment of missense mutations in the *TP53* DBD (17, 18), while revealing differences in mutation types and locations between myeloid neoplasms, including relative enrichment of null and splice-site mutations in AML-MRC compared to MDS.

**Figure 1:**
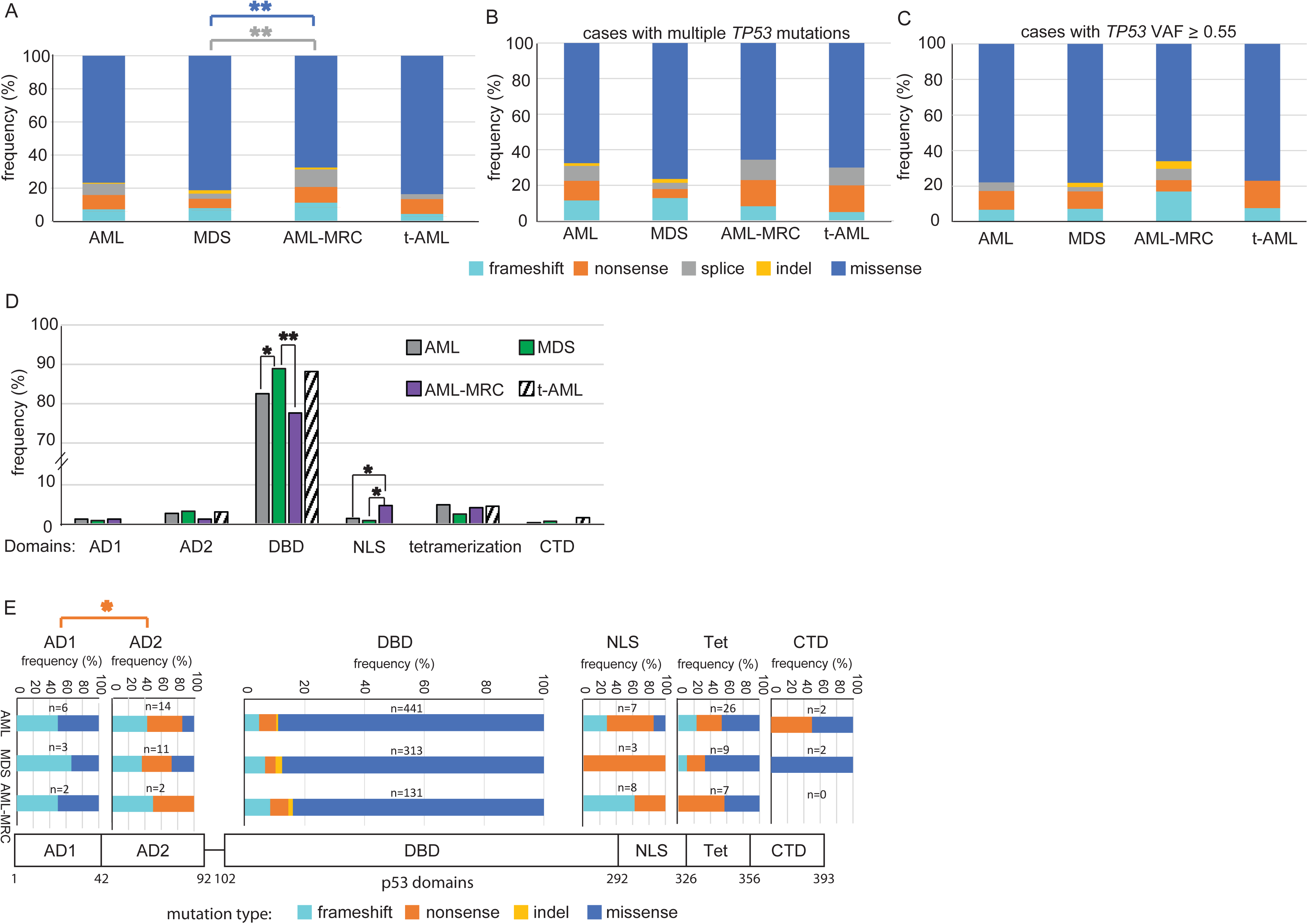
*TP53* mutation types and locations differ between myeloid neoplasms. A) Frequency of frameshift, nonsense, indel, splice, and missense *TP53* mutations shown as a percentage of total *TP53* mutations in the indicated disease categories. Curved brackets indicate null mutations. B) Frequency of each mutation type in cases with two or more *TP53* mutations in the indicated neoplasms (number of mutations: AML n=129, MDS n=107, AML-MRC n=40, t- AML n=14). C) The frequency of each type of mutation was plotted as the percentage of total mutations of VAF ≥ 0.55 in the indicated diseases. AML n=103, MDS n=41, AML-MRC n=47, t-AML n=13. D) Frequency of mutations in each p53 domain shown as percentage of total number of mutations in each disease category. (A,D) AML n=536, MDS n=352, AML-MRC n=167, t-AML n=65. E) Frequency of each mutation type by domain in AML, MDS, and AML- MRC. Domain boundaries are indicated. For adjacent domains, the C-terminal end of the boundary is shown. The total number of mutations in each domain is shown for each disease. Splice mutations were excluded in this analysis. * p <0.05, ** p <0.01 by Chi-squared test with Bonferroni correction.

### Analysis of distribution of *TP53* mutations reveals myeloid-specific and disease-specific hotspots

To obtain a more granular view of the types and locations of *TP53* mutations, we constructed lollipop plots depicting the frequency of missense, nonsense, and frameshift mutations at each codon in each disease (**Fig. 2A**). 195 out of 393 amino acids were mutated at least once. 128 mutated positions harbored missense mutations, and 27 positions showed examples of both missense and null mutations, with a general bias towards missense mutations. To determine whether the overall pattern of mutations (their locations and frequencies) differed between neoplasms, we plotted the cumulative fraction of all mutations along the length of the protein (**Fig. 2B**). The greatest differences in the cumulative curves were seen between MDS and AML-MRC, with AML-MRC being biased towards C-terminal mutations (**Fig. 2B**). To identify individual hotspot mutations in a quantitative and unbiased manner, we applied an algorithm that detects peaks in mutation frequency above local background levels. This analysis was conducted only for missense mutations due to the relative paucity of other mutation types. Of the seven most common *TP53* mutation sites in human cancers (14) (subsequently referred to as *pan-cancer hotspots*), six were identified as peaks in AML(R175, Y220, G245, R248, R273, and R282); R249 was the only pan-cancer hotspot not identified as such in myeloid neoplasms (**Fig. 2C**). MDS and AML-MRC additionally lacked mutations at G245 and R282. Hotspot mutations have been classified as “contact” or “conformational” mutations based on their effects on DNA binding or protein folding, respectively (19). Contact mutations (R248Q, R273H, and R282W) were more common in AML-MRC (16.8%) than in either AML or MDS (10.9 and 12.9%, respectively), whereas conformational mutations (R175H, Y220C, G245S, R249S) occurred with nearly equal prevalence in all categories (11.4-12.4%). In addition to the 7 pan-cancer hotspots, we identified several hotspots present in 2 of 3 diseases, which we termed “myeloid-specific” (irrespective of their mutation rates in individual solid tumors). “Disease-specific” hotspots were defined as being present in only one disease. These results highlight the overall similarities in mutation patterns between myeloid neoplasms while also revealing differences in mutation distributions (frequency and location).

**Figure 2:**
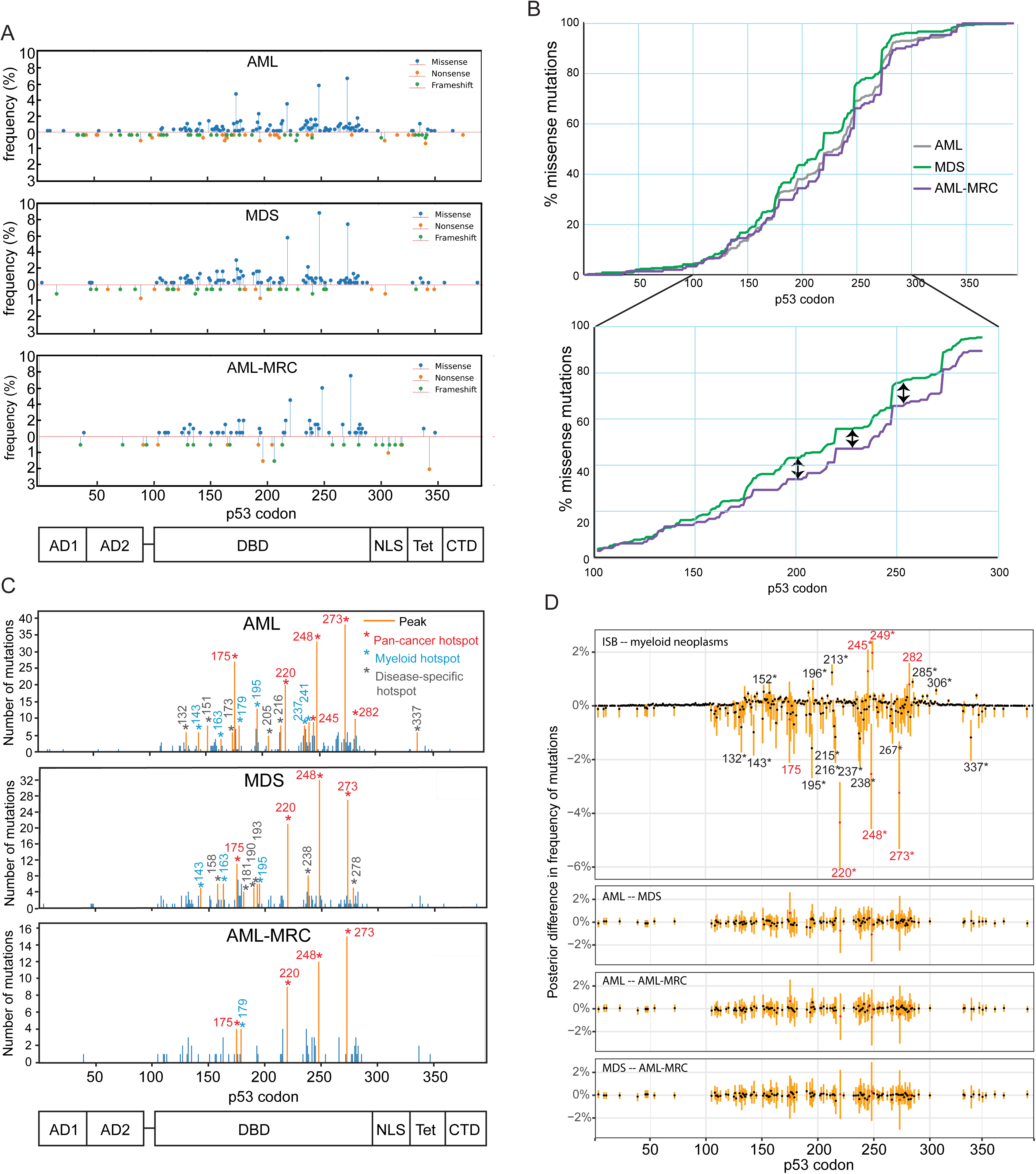
TP53 mutational hotspots in myeloid neoplasms overlap with solid tumors, but also include myeloid- and disease-specific peaks. A) Lollipop plot showing the frequency of missense (above axis) and nonsense and frameshift mutations (below axis) in the indicated disease categories. Frequency is shown as a percentage of total *TP53* mutations in each disease category. AML n=496, MDS n=334, AML-MRC n=147. B) Cumulative curve depicting the fraction of *TP53* missense mutations as a function of position (codon number) within the protein. Inset shows the cumulative curve in the DBD region for MDS and AML-MRC. Arrows show examples of regions where the AML-MRC curve lags the MDS curve. C) Identification of missense hotspots in the indicated diseases by peak-finding algorithm. Orange lines represent peaks identified by the algorithm, blue lines represent mutation sites not identified as peaks. Hotspots are classified as pan-cancer (from (14)), myeloid-specific (occurring in 2 or more myeloid categories in this dataset), or disease-specific (unique in this dataset). D) Differences in underlying mutation frequency between the indicated disease categories at *TP53* codons with missense mutations were inferred using data from Fig 1A. Black points represent the expected value and orange lines the central 95% Bayesian credible interval. Codons with no recorded mutations in the compared disease groups (ISB and myeloid neoplasms; and AML, MDS, and AML-MRC) were not considered. The expected difference in underlying mutation frequencies of hotspot codons are colored red. (B-D) AML n=411, MDS n=286, AML-MRC n=113.

We next performed a Bayesian analysis to determine to what degree codons with missense mutations differed in their underlying mutation frequency between diseases. This analysis was performed for the pooled dataset of myeloid neoplasms compared to the ISB-CGC dataset of mutations derived from all cancer types to assess differences between hematologic malignancies and solid tumors (20). It was separately conducted pairwise for the different categories of myeloid neoplasms (**Fig. 2D**). By sampling from the distributions of inferred mutation frequencies and then the difference between them, this process yields a simulated expected difference with 95% Bayesian credible interval for each codon. The largest expected difference between myeloid neoplasms and the pan-cancer dataset was observed for mutations at codon Y220, which was enriched in myeloid neoplasms; mutations at R175, R248 and R273 were also over-represented in myeloid neoplasms, while mutations at R245, R249, and R282 were under-represented. (**Fig. 2D**, top panel). Estimated differences between the categories of myeloid neoplasms were smaller than those between myeloid neoplasms and solid tumors, with no individual codons at which the 95% Bayesian credible interval of the difference in mutation frequency excluded zero in any of the pairwise comparisons (**Fig. 2D**, bottom).

We next catalogued the amino acid changes resulting from missense mutations in each disease. 77 codons showed at least two different substitutions (shown in **Fig. 3**). C176 had the greatest diversity of substitutions, with mutations to F, G, R, S, W, Y detected across diseases. The frequencies of specific substitutions varied between diseases at some positions. R273H was more prevalent in AML-MRC than in AML (9.5% vs 5.8%), with MDS showing an intermediate prevalence (7.3%) (**Fig. 3**). R248 was biased towards substitution to Q over W in MDS, whereas AML-MRC showed the opposite trend, and AML showed almost no bias. These patterns suggest both flexibility and specificity in pathogenic *TP53* mutations.

**Figure 3:**
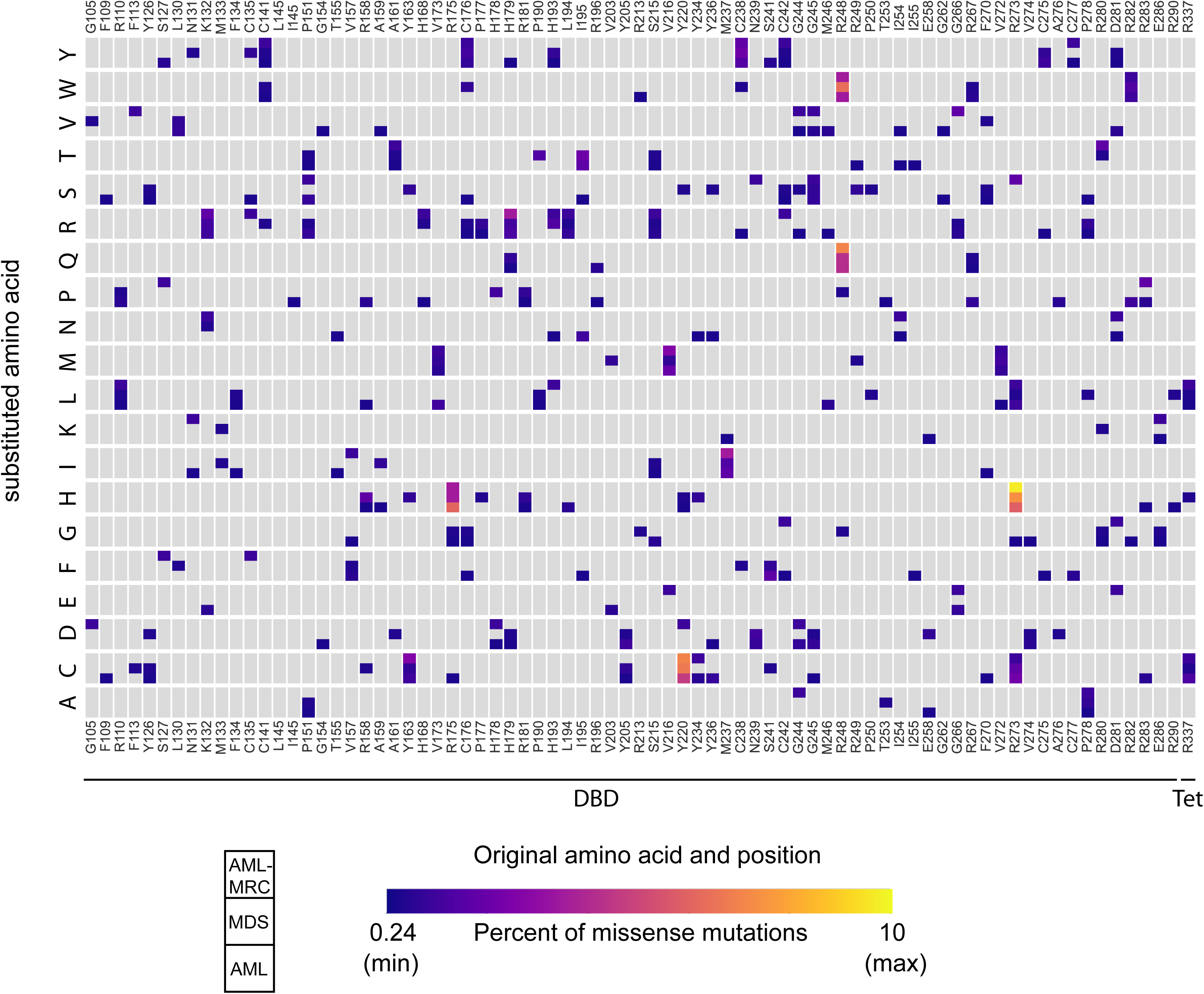
*TP53* missense mutations show a wide diversity and disease-specific biases in amino acid substitutions. Heat map depicting the frequency of each type of amino acid substitution at each codon. Only codons displaying multiple types of substitutions are shown. Each cell depicts the frequency of substitution in each of the 3 disease categories as depicted in the key. Grey boxes indicate no substitutions detected. The left end of the color spectrum represents one mutation event in the largest dataset (AML). AML n=411, MDS n=286, AML- MRC n=113.

Overall, our findings demonstrate a distinct distribution of p53 mutations in myeloid neoplasms compared to solid tumors, and suggest distinct mutational patterns between individual myeloid neoplasms. The differences in amino acid substitutions between diseases at particular codons suggest distinct functional roles for specific p53 mutations in disease initiation and progression.

### Functional analyses of *TP53* mutations

We assessed the functional consequences of missense p53 mutations by calculating the distribution of transcriptional activity scores in each neoplasm (15) (see Methods), and found that it was significantly higher in MDS compared to AML-MRC, with AML falling between the two (**Fig. 4A**). This result held true when the analysis was performed only on the subset of cases with VAF ≥ 0.55, indicating that it was independent of the status of the second allele (**Fig. 4B**).

**Figure 4.**
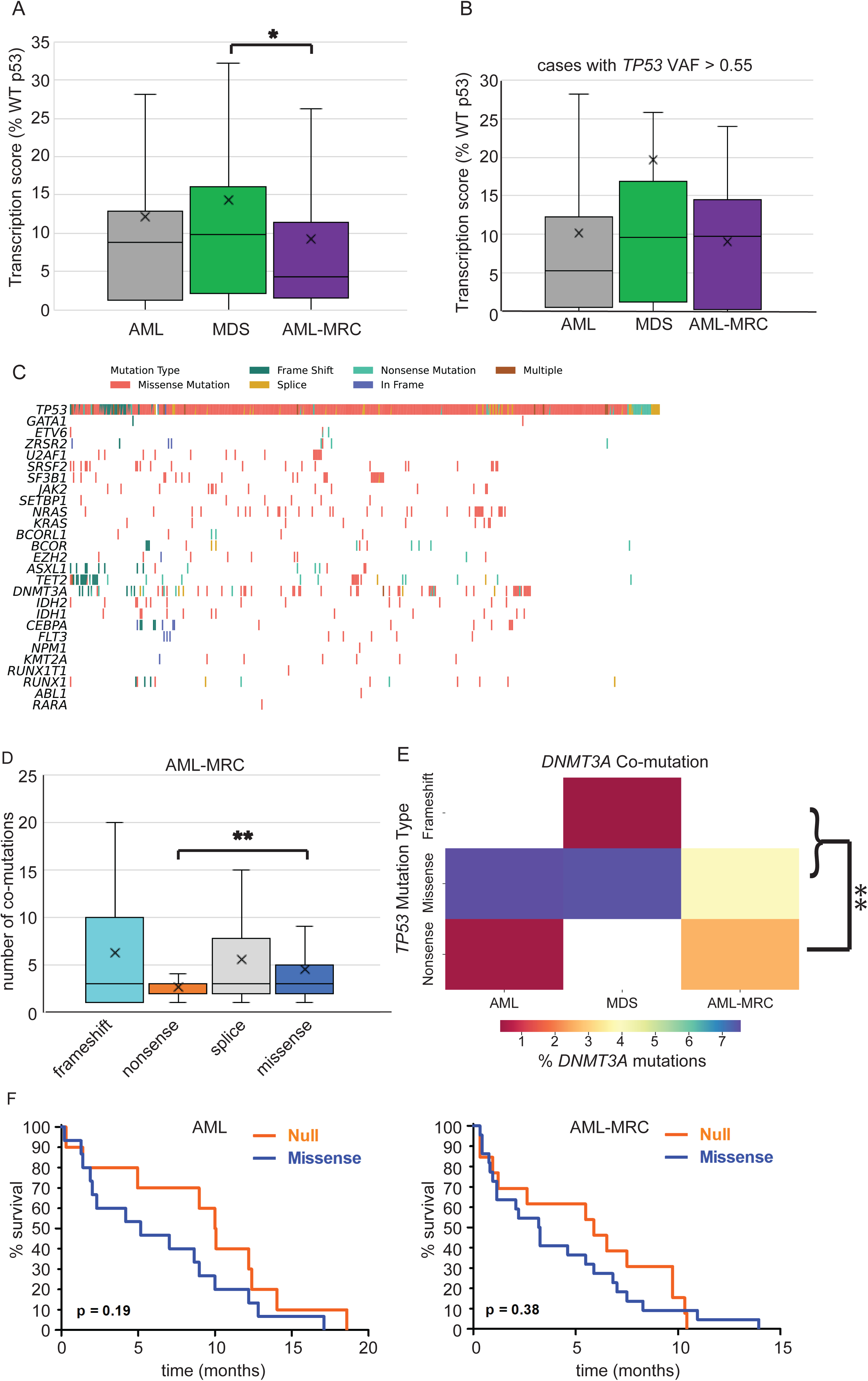
p53 transcriptional activity and co-mutations vary between classes of neoplasia and *TP53* mutation types. A) Box and whisker plots of transcriptional activity scores of missense mutants in the indicated disease categories. AML n=408, MDS n=284, AML-MRC n=113. * p< 0.05 by Welch’s t-test with Bonferroni correction. B) Box and whisker plots of transcriptional scores of missense mutants with VAF ≥ 0.55 in the indicated neoplasms. AML n=79, MDS n=32, AML-MRC n=30. A-B) Horizontal line indicates median, X marks the mean, and box top and bottom represent the 75th and 25th percentiles, respectively. Error bars represent the 95% confidence interval. C) Co-mutations in *TP53* with the indicated leukemic driver genes, segregated by *TP53* mutation type. D) Box and whisker plots of the number of co-mutations occurring with each type of *TP53* mutation in AML-MRC (n=195). ** p< 0.01 by Welch’s t-test with Bonferroni correction. E) Co-mutations of *DNMT3A* with the indicated classes of *TP53* mutations in AML, MDS, or AML-MRC. Color indicates the percent of *DNMT3A* mutations co- occurring with the indicated class of *TP53* mutation. ** p< 0.01 by chi-squared test comparing the association of *DNMT3A* mutation with the indicated *TP53* mutation type vs. all other *TP53* mutation types. F) Survival curves of AML (n=10 null and 15 missense *TP53*) or AML-MRC (n= 13 null and 22 missense *TP53*) patients with the indicated *TP53* mutation types. p-values calculated by Gehan-Breslow-Wilcoxon test.

To gain further insight into the interactions of mutant *TP53* with other oncogenic pathways, we investigated patterns of co-mutation between *TP53* and 24 known leukemic driver genes. *DNMT3A*, *TET2*, and *NRAS* were commonly co-mutated with *TP53,* while *FLT3* and *NPM1* co-mutations were significantly less common than in cases with WT *TP53*, consistent with previous studies (16, 21, 22). When parsed by *TP53* mutation type, co-mutations in the 24 driver genes were under-represented in *TP53* nonsense and splice mutations (**Fig. 4C**): across samples with both single and multiple *TP53* mutations, 39% and 59% of samples with nonsense and splice mutations harbored at least one co-mutation, respectively, compared to a co-mutation rate of 88% for *TP53* missense mutations. When considering the total number of co-mutations (in addition to the selected 24 driver genes), AML-MRC with nonsense *TP53* mutations showed significantly fewer co-mutations compared to other *TP53* mutation types (**Fig. 4D**). However, in AML-MRC, nonsense *TP53* mutations were significantly associated with co-mutation of *DNMT3A* (**Fig. 4E**), with 4 of 10 *DNMT3A* mutations occurring with nonsense *TP53* (for comparison, no *DNMT3A* mutations (of 22) in MDS co-occurred with nonsense *TP53*). These findings provide further evidence that *TP53* null and splice-site mutations may mediate pathogenesis of myeloid neoplasms through distinct mechanisms from those of the more common missense mutations.

### *TP53* null mutations tend to show longer survival in AML and AML-MRC

To investigate whether *TP53* mutation types affect patient outcomes, we first grouped *TP53* mutations as missense or null, with the latter category comprised of frameshift, nonsense, and splice mutations. This grouping was chosen because of the distinct effects of the mutation type on p53 protein: null type mutations do not express detectable protein, whereas missense mutants tend to overexpress it. Although not statistically significant, in both AML and AML-MRC, we detected longer survival in p53-null cases, albeit shorter than the survival reported for WT p53 (10, 13) (**Fig. 4F**). These results suggest that *TP53* missense mutations may exert oncogenic effects in AML and AML-MRC beyond simply inactivating its tumor-suppressive function.

## Discussion

In this study, we analyzed *TP53* mutations in myeloid neoplasms, and identified differences in *TP53* mutations between myeloid neoplasms and solid tumors as well as between subtypes of myeloid neoplasms, particularly between myelodysplastic syndrome and acute myeloid leukemia with myelodysplasia-related changes. Our findings suggest discrete biological mechanisms for mutant p53 in hematologic malignancies, and particularly in driving initiation and progression of myeloid neoplasia, and indicate that molecular characterization of *TP53* mutations, as opposed to p53 immunohistochemistry alone, is required for optimal assessment of *TP53*-mutant myeloid neoplasms in clinical practice.

We identified significant differences in *TP53* mutations between myeloid neoplasms and solid tumors. Most notably, we observed marked enrichment of mutations at codon Y220, which are relatively uncommon in solid tumors but were among the most frequent mutations in myeloid neoplasms in our dataset. This finding is of particular interest given that this mutant protein is uniquely susceptible to reactivation of small-molecule stabilizers (23); such molecules are currently being evaluated in clinical trials for solid tumors (e.g., NCT04585750) (24). Our findings argue for prioritizing evaluation of these agents in myeloid neoplasms, and illustrate the potential benefit of systematically identifying *TP53* mutations in myeloid neoplasms to identify patients who might benefit from this and other mutation-specific therapies.

Among myeloid neoplasms, MDS and AML-MRC showed the greatest differences in terms of their types, distributions, and predicted transcriptional activities of *TP53* mutations. These findings suggest that distinct types of *TP53* mutations mediate initiation of MDS and progression of MDS to AML. We find that R175H, G245S, and R282W were under-represented in MDS and AML-MRC compared to AML; the R175H result is consistent with the study of Tashakori *et al* (16). Conversely, R273H mutations were more common in MDS and AML-MRC than in AML. Our finding that missense mutations, particularly those with higher levels of retained transcriptional activity, are over-represented in MDS, whereas mutations resulting in complete loss of expression or transcriptional activity of p53 protein are relatively over-represented in AML-MRC, suggests that p53 missense mutations may play a more active role in initiation of MDS, possibly by acquiring gain-of-function properties, whereas p53 inactivation is sufficient for progression to AML-MRC. The specific association of *TP53* nonsense mutations with *DNMT3A* mutations in AML-MRC supports this hypothesis, since *DNMT3A* mutations (and, more broadly, mutations in genes that regulate chromatin accessibility and/or epigenetic control of gene expression) are common “founder” mutations in both MDS (4) and clonal hematopoiesis of indeterminate potential (CHIP) (25). Overall, these findings raise the intriguing possibility that specific identification of TP53 mutations in MDS may be useful in predicting which patients are likely to progress to AML. Tracking the evolution of TP53 mutations in individual patients during progression from MDS to AML-MRC will be instrumental in further evaluation of this question.

In both AML and AML-MRC, inactivating mutations tended to show longer survival than missense mutations. These findings contrast with a recent report (16) that found no survival difference between *TP53* mutation types; however, this study did not distinguish between categories of myeloid neoplasia. Our data suggest that more detailed multivariate analyses that account for neoplasm type and for *TP53* mutation type will be required to conclusively determine the effects of discrete classes of *TP53* mutations on patient survival. Notably, any differences in survival could arise from differences in *TP53* mutations themselves or from the spectrum of co-mutations, which varied according to neoplasm subtype as well as *TP53* mutation types. Specifically, *DNMT3A* mutations were associated with AML-MRC containing nonsense *TP53* mutations, but the consequences of this association remain unknown.

Clinical assessment of *TP53* mutation has historically operated under the guiding assumption that loss of functional p53 protein activity mediates its role in driving neoplasia, and, therefore, that specific identification and characterization of mutations is not clinically useful, and that p53 immunohistochemistry (a proxy for loss of protein function) is sufficient for diagnostic workup. This assumption is embedded in recent updates to classification schema for hematologic malignancies; both the 5th edition World Health Organization classification and the International Consensus Classification of myeloid neoplasms incorporate *TP53* mutation status into classification of myeloid malignancies (11, 12), but neither scheme incorporates mutation type or functional consequence. Our findings suggest that such assessment may be incomplete, and that\ assessment of *TP53* mutation type and consequences may be necessary to fully account for its impact on disease biology and prognosis.

## Methods

### Study cohort

We identified *TP53* mutant cases of AML, MDS, AML-MRC, and t-AML in the cBioPortal database, a public-facing repository that incorporates data from multiple studies (26, 27), and in the Hematologic Malignancies Data Repository (HMDR) of the Dana-Farber Cancer Institute. All cases in both datasets were annotated with diagnostic classification and mutation(s) identified by next-generation sequencing, including nucleotide change, amino acid change, and variant allele fraction (VAF); additional information, including karyotype, treatment history, and clinical outcome, was available for a limited subset of cases. Duplicates within and between databases were manually filtered and removed, and only cases with *TP53* mutations annotated as pathogenic or likely pathogenic were included in the analysis (i.e., non-pathogenic variants were excluded). After filtering, there were a total of 1,120 independent instances of *TP53* mutations in myeloid neoplasms, representing 536, 352, 167, and 65 instances in AML, MDS, AML-MRC, and t-AML, respectively. We classified *TP53* mutations as nonsense, frameshift, splice, in-frame indel, or missense, and calculated the frequency of each mutation type in each disease.

Information on large deletions was not available. When the same patient was sequenced multiple times, replicates were removed for cataloguing p53 mutations.

### Peak Finding

Peaks in mutation frequency above background were identified by find_peaks from SciPy’s Signal processing in Python 3.8.

### Co-mutation analysis

Data were obtained from cBioPortal using the cBioPortalData Bioconductor package in R 4.1.3 from the previously identified studies. All co-mutation plots were created using the CoMut python package.

### Statistical analyses

Continuous variables were compared by Welch’s t-test. Categorical variables were compared pair-wise by Chi-Squared tests. Fisher tests were used for co-mutation studies. In all cases, Bonferroni corrections for multiple hypothesis tests were applied.

### Comparison of underlying mutation frequencies across disease categories

Using the dataset of missense mutations in AML, MDS, and AML-MRC, letting *d* denote the disease and *l* the number of codons, we observed a vector of integers *y*_*d*_ = (*y*_*d*1_, *y*_*d*2_, …, *y*_*dl*_) for each disease. 411 mutations were observed among AML patients, 286 for MDS, and 113 for AML-MRC. Considering only codons with at least one mutation recorded under any disease, we have that *l* = 121. Assuming *y*_*d*_ ∼ Multinomial(∑*y*_*d*_, *θ*_*d*_) and 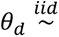 Dirichlet(*α*), then *θ*_*d*_|*y*_*d*_ ∼ Dirichlet(*α* + *y*_*d*_) . We set *α* such that *α*_*i*_ = ∑_*d*_ *y*_*di*_,, as an empirical prior. We compared myeloid neoplasms with the ISB-CGC dataset using a similar framework, but with different priors and *l* = 378. Specifically, we set *θ*_ISB_ to be Dirichlet-distributed with concentration parameter 0.1, and *θ*_MN_ ∼ Dirichlet(*y*_ISB_/∑*y*_ISB_ × 35). Code used for sampling can be found at https://github.com/berkalpay/tp53mutations/.

### Transcriptional activity analysis

Transcriptional activity scores were obtained from the PHANTM database (http://mutantp53.broadinstitute.org), which draws its data from the study of Kato *et al*.(15)

## Data Availability

All data produced in the present study are available upon reasonable request to the corresponding author (please contact). Code used for statistical and computational analysis can be found at https://github.com/berkalpay/tp53mutations/.

https://www.cbioportal.org/

https://github.com/berkalpay/tp53mutations/

## Data sharing statement

For original data, please contact.

## Acknowledgments

These studies were supported by funding from the Ludwig Institute at Harvard (to A.J. and G.L.), NIH R35 GM139572 (to G. L.), and the National Science Foundation Graduate Research Fellowship under Grant No. DGE-2140743 (to B.A.A.). Some results were presented as a selected oral abstract at the United States and Canadian Academy of Pathology (USCAP) annual meeting on March 14, 2023. The authors would like to thank Geoffrey Fell (Dana-Farber Cancer Institute), Derek Aguiar (University of Connecticut), Eliot Fenton (Harvard University), and Marie-Abele Bind (Massachusetts General Hospital) for help with statistical analysis; Jon Aster, Scott Rodig, and Annette Kim (Brigham and Women’s Hospital) and Sandy Nandagopal and Meg Dillingham-McCullough (Harvard Medical School) for advice and critical review of the manuscript; and the members of the Brigham and Women’s Hospital Department of Pathology and Harvard Medical School Department of Systems Biology for helpful discussion.

## Authorship Contributions

A.J. and S.B.L. designed the concept, developed the study, and wrote the original draft of the manuscript. E.E.A. and B.A.A. performed computational and statistical analysis of data. G.L. advised the study and provided essential resources. All authors contributed to the final version of the manuscript.

